# The prevalence of neuropathic pain pathophysiology associated with ankle fracture: A study protocol

**DOI:** 10.1101/2025.06.19.25329965

**Authors:** Tyler Nguyen, Kelly M. Naugle, Michael Fletcher, Hillarie Dawn Arellano, Kathy Leslie, Anastasiya Bahdanovich, MaKenzie Barger, Lauren Christine Hill, Roman M. Natoli, Fletcher A. White

## Abstract

Chronic pain is prevalent among U.S. military personnel and often accompanied by comorbid behavioral health disorders and other medical conditions that further complicate its management. According to the Centers for Disease Control and Prevention, the prevalence of chronic pain among active-duty Service members is 1.5 to 2 times higher than the 20% of American adults who live with chronic pain. Recent report findings determined that Service members make up a large population within the Military Health Systems (MHS), and that this population is disproportionately affected by lost duty days, early retirement, loss of readiness, and increased burden to the MHS. To date, the Department of Defense (DOD) and MHS have emphasized multimodal, multidisciplinary, stepped treatment for chronic pain that prioritizes nonpharmacologic therapies and non-opioid pain medications. Though the DOD and MHS have invested in several pain treatment types, our level of understanding needs to better distinguish between acute and chronic pain and identify risk factors and mechanisms responsible for the chronification of pain, as it is the chronic pain which compromises functioning and readiness to a greater degree across the force. The novel information generated by this study will enhance our understanding of how ankle fracture elicits pathological risk factors for bone fracture associated neuropathic pain (BFNP), which ultimately impairs health-related quality of life. Due to the high prevalence of ankle fractures and the subsequent risk of developing chronic pain after ankle fracture, we will utilize this patient population to provide the preliminary evidence on whether bone fracture and subsequent BFNP phenotypes are reflected in specific genetic profiles and activated states of immune cells.

## Introduction

Persistent pain, such as neuropathic pain (NP), is an outcome of ankle fracture repair with incidence rates 1-year post-surgery of 18-42% [1]. Ankle fractures are one of the most common surgically-treated fractures in adults with the greatest incidence occurring in young males [2]. In general, three stages of pain follow a bone fracture including acute, subacute and chronic pain. Acute pain occurs immediately after injury. After about a week or two, the worst pain is usually subsiding as the fractured bone and the surrounding soft tissue begin to heal. Many individuals may continue to report pain during this subacute period. Pain that persists after fracture union has taken place is called chronic pain. Chronic or persistent NP is one of the worst, longest-lasting, and difficult symptoms to manage after fracture repair in civilian and military populations [3]. Some of the pathophysiologic mechanisms leading to NP likely propagate early after injury, providing opportunity for early interventions.

Neuropathic pain-associated with bone fracture likely originates from a lesion affecting the somatosensory nervous system and may be associated with abnormal sensations, called dysesthesia, or pain from normally non-painful stimuli (allodynia). The condition may have continuous and/or episodic (paroxysmal) components, with the latter resembling stabbing pain or electric shocks. The condition of NP also tends to affect defined dermatomes and there may be limits to the area of pain. The general working principle is that the lesion leading to pain must directly involve nociceptive pathways [4]. Additional elements which can contribute to NP include sensitization of intact nociceptors that survive injury, which innervate the region affected by the injured nerve fibers and contribute to spontaneous activity. These changes in the intact nociceptors may induce ongoing pain and account for certain aspects of hyperalgesia [5]. Conditions associated with bone fracture-associated NP (BFNP) include traction neuropathy, nerve compression from soft tissue edema, bone fragment, implants, and hematoma [6, 7].

In civilian adult populations, prevalence rates of NP are about 1 in every 10 adults over age 30, though the prevalence rate and people identified vary depending on the method of identification of NP [8, 9]. Given the number of active duty personnel and Veterans who experience pain due to injury, the US military instituted a number of programs, guidelines and initiatives to better manage acute pain for combat-related injuries [10]. Despite the instituted practices by the military, NP after fracture remains a major problem, and literature documenting detailed outcomes of BFNP is scarce.

Chronic bone injury sequelae, which may contribute to BFNP, include the release of factors that excite and sensitize sensory nerves including neurotransmitters, cytokines/growth factors and receptors, upregulation of sensory neuron ion channels, induction of nerve fiber sprouting in bone tissue, and central sensitization in the brain which can amplify pain signals [11–14]. More recent computational approaches suggest that it is also possible that networks of biomarkers exist that could accurately identify individuals at high risk of BFNP. These biomarkers may exist at the time of injury and are likely accessible at early time points in fracture care. Changing the standard of care treatment for BFNP by identifying at-risk patients early after injury could lead to decreased economic burden in treating BFNP and mitigation of the substantial decrease in quality of life these patients experience.

The primary aims of this longitudinal study are: (1) To evaluate how dysfunctional endogenous pain modulation and increased pain sensitivity will be associated with increased risk for development of BFNP and (2) to identify peripheral blood immune cell biomarkers for BFNP. The hypothesis of this study is that bone fracture will induce dysfunctional endogenous modulation of pain (i.e., increased peripheral and central sensitization) and a long-term systemic alteration of the innate immune system, both of which may increase the risk for NP in bone fracture patients who develop BFNP compared to bone fracture patients who do not develop chronic pain and healthy controls. Our heuristic model linking bone fracture to neuropathic pain (Fig 1). Once these differences are quantified and biological signaling pathway analyzed, we will be able to identify a biosignature, or set of biomarkers, that can be used to identify BFNP susceptibility early after fracture.

**Fig 1.**
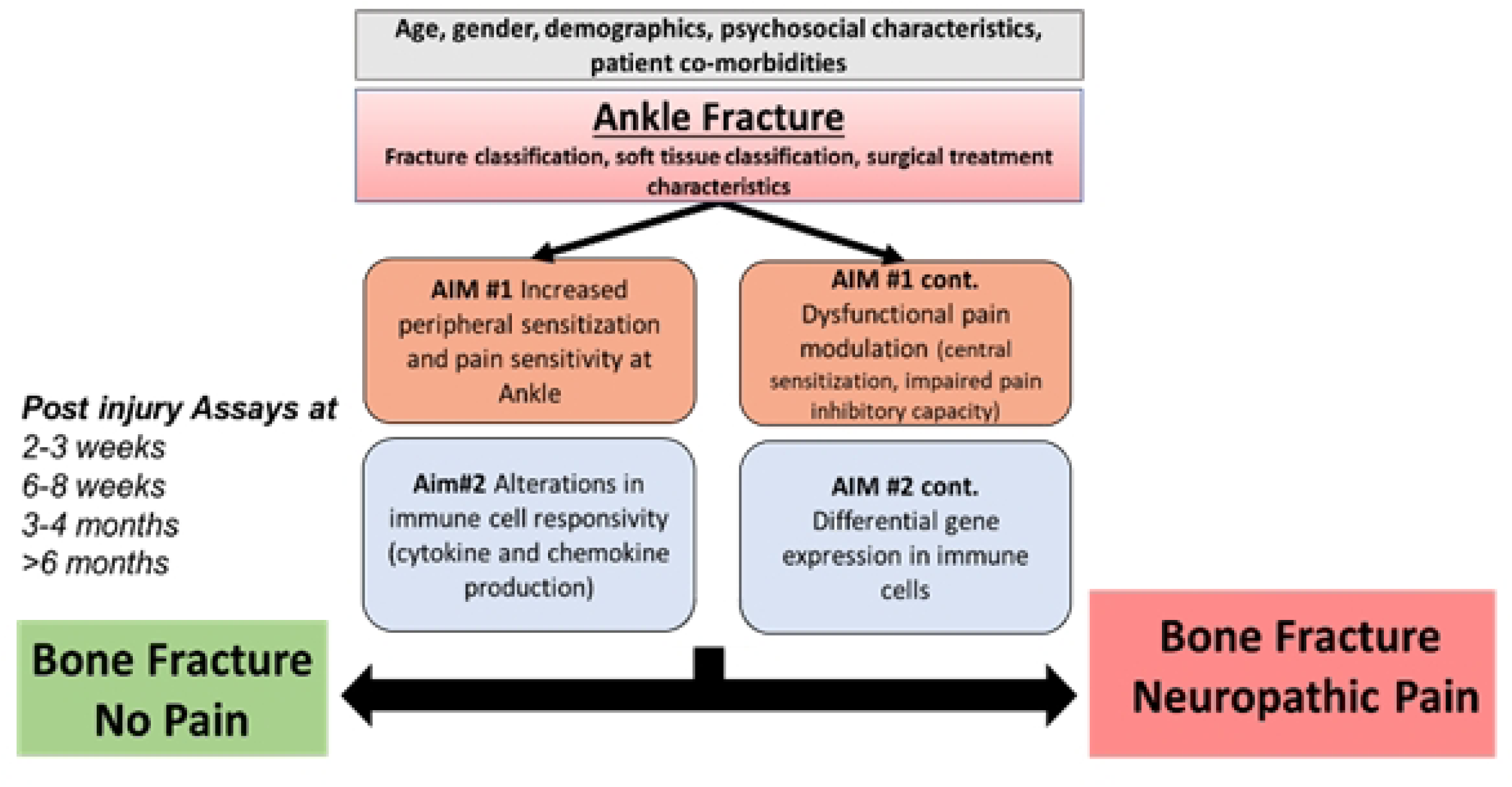
Heuristic model linking bone fracture with chronic neuropathic pain.

## Methods

### Study design and setting

This study will utilize a prospective cohort study design (Fig 2). We will enroll participants who have sustained rotational ankle (specifically AO/OTA 44 types A-C [15]) fractures that are treated operatively. Fracture patients will complete clinic assessment sessions at standard of care visits, including their first post-op appointment (2-4 weeks), at 5-9 weeks, at 3-5 months post-operatively, and at a 6-9 months follow-up visit. Clinic assessment sessions will include completion of questionnaires, blood draw, thermal imaging, quantitative sensory testing, and clinical assessment of infection, fracture healing, and radiographic arthritis. Questionnaires will be completed over the phone or in- person at each approximate time point. These visits will allow assessments during the subacute and chronic stages of BFNP. If fracture participants do not show for the in-person standard of care 6-month visit, the participant will be contacted by phone to collect patient reported outcomes (PROs). Along with this, at 1-1.5 years out the patient may be called for self-reported outcomes to assess if patients reporting chronic pain at 6-9 months still have persistent pain. A separate group of 40 uninjured healthy control participants will perform the same assessments during one study session. Enrollment procedures, including assessment of inclusion/exclusion criteria and informed consent, will occur in study session 1. This study was approved by the Indiana University (IU) Institutional Review Board. The study began January 1^st^, 2024 and will be completed on December 31^st^, 2028. We currently have 63 clinical subjects to date and expect to complete enrollment in June 2028. Data collection and results will be completed in 2029. No analysis has been implemented at this time.

**Fig 2.**
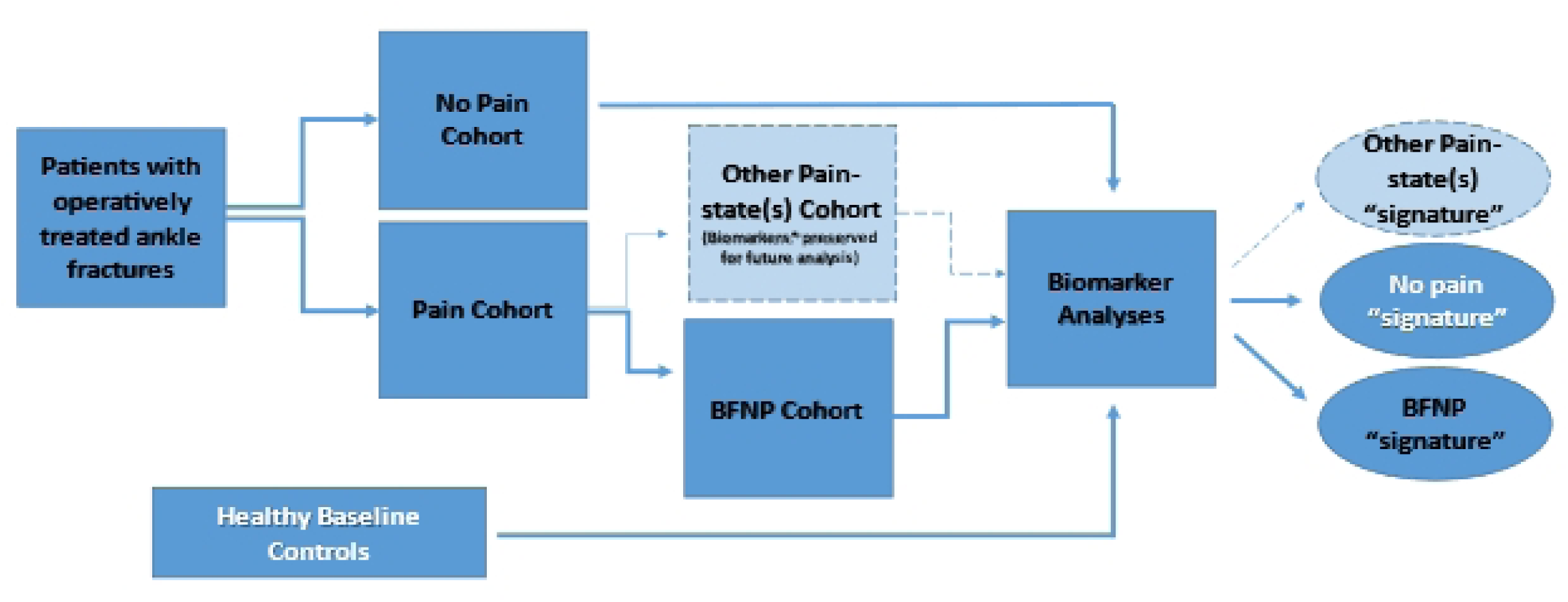
Prospective Cohort Study Design.

#### Study design

This study is an observational cohort study of patients with rotational ankle fractures. Patients will be followed longitudinally, with assignment of patients into 3 groups at >6 months after surgery for fracture repair. The distinction between the *Pain* and *No Pain* cohort will be based on the Chronic Pain Grading Scale (CPGS), with a CPGS of 0 at >6 months defining *No Pain* and a CPGS ≥1 defining the *Pain* Cohort. Several pain states may exist in the *Pain* Cohort. The Douleur Neuropathique 4 (DN4) will be used to identify patients with *BFNP* as described in the text. Patients not meeting *BFNP* will be placed in the *Other Pain-state* group (e.g., CRPS, inflammatory pain, arthritis, pain from complications). This group will be further divided into patients with CRPS and those without based on the CRPS Research Criteria. This study will focus on identifying early biomarker signatures that can discriminate between *No Pain* and *BFNP*. Biomarkers include quantitative sensory testing (QST), mRNA expressions and immune cell activation, and thermography.

### Participants

#### Sample size

For fracture patients, we will enroll 265 participants who have sustained rotational ankle (specifically AO/OTA 44 types A-C [15]) fractures that are treated operatively. For control patients, we aim to enroll up to 40 healthy adults, who have never sustained a bone fracture. We will attempt to enroll the same percentage of males and females as the fracture patients.

The sample size was calculated based on the limited data from literature and our experiences on prior studies [16–18]. Dietz et al. reported that patients with fracture or trauma that developed complex regional pain syndrome (CRPS) had higher scores for pain, disability and all patient-reported outcomes than fracture controls [18]. Specifically, the effect sizes of the differences between these two groups were calculated as: Cohen’s d=.31 for neuropathic pain symptom inventory (NPSI), d=.34 for current pain, and odds ratio=7.4 of developing allodynia. Hall et al. reported the percentage of Brief Pain Inventory (BPI - today’s pain) as 55%, 58% and 34% in the 1st, 2nd and 6th months, respectively, among 53 limb fracture patients [19]. In addition, Rbia et al. [7] and Friesgaard et al. [19] reported that about one in four ankle fracture patients would develop persistent neuropathic pain. Data on other outcomes are sparse. Based on the above information and assuming type one error of α=0.05 and correlation between repeated measures as ρ=0.6 and ratio for group size as 1:4, we estimated the total sample size needed to achieve sufficient power (β≥0.8) were n=205 for NPSI, n=170 for current pain and for allodynia, and n=213 for BPI. To maximize our ability to detect the differences, we chose the largest sample size n=213. Our experiences following fracture patients longitudinally revealed 17% attrition rate (internal surveillance data for PREP-IT studies) [17, 20]. We conservatively assumed the overall attrition rate to be 20%. Therefore, a total sample of N=265 (expected to generate 53 BFNP patients) would need to be recruited.

#### Inclusion/Exclusion criteria

All fracture patients must meet all inclusion criteria and be absent of all exclusion criteria to be enrolled in the study. The criteria are listed in Table 1.

**Table 1.**
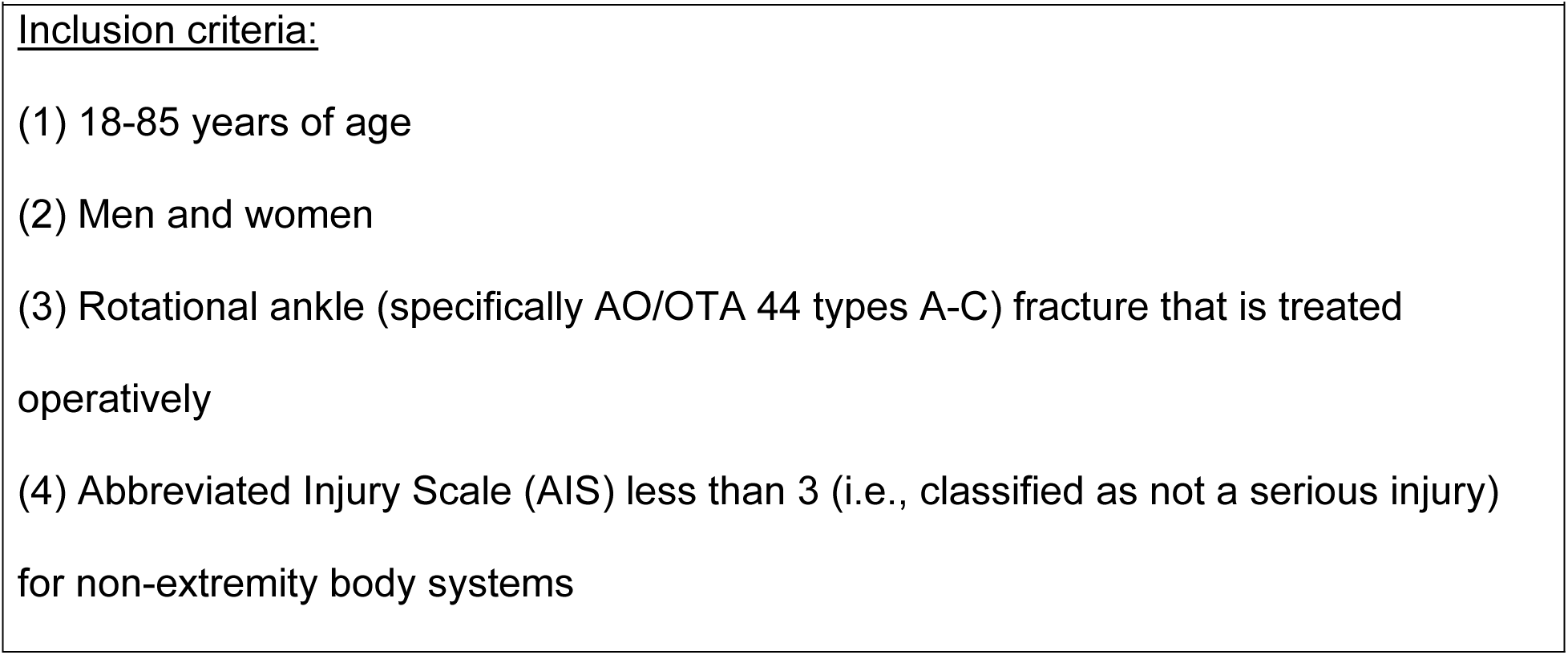
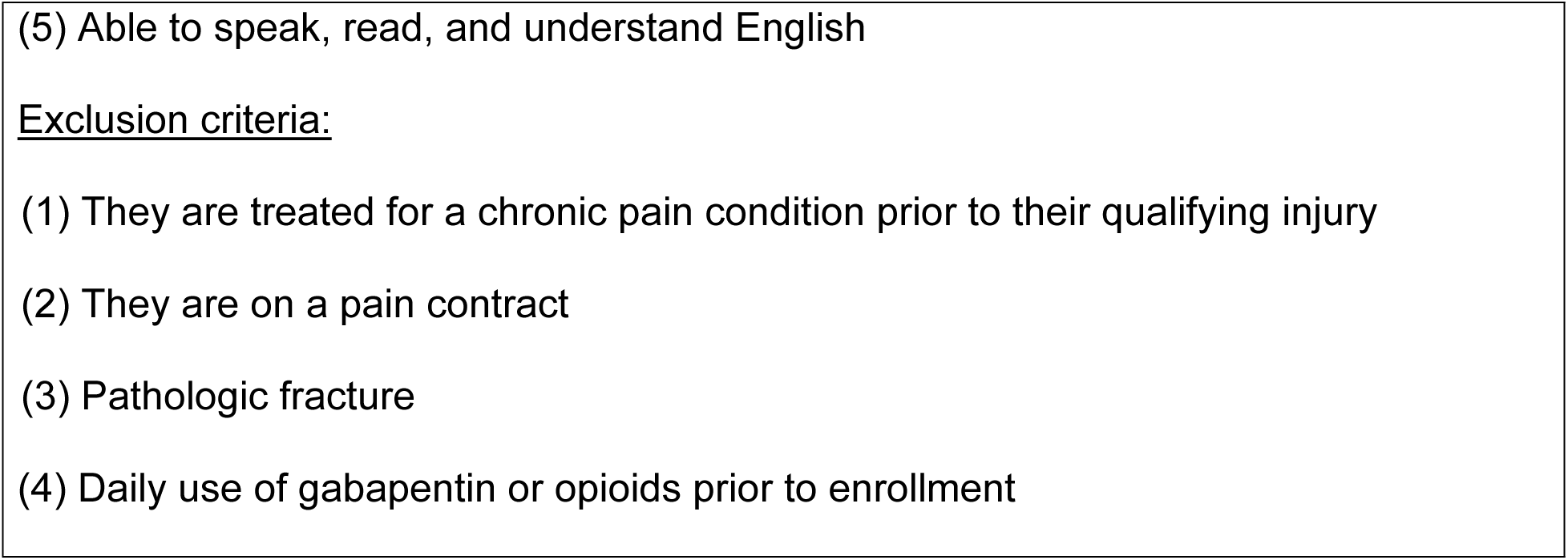
Inclusion and exclusion criteria for fracture patients.

All control patients must meet all inclusion criteria and be absent of all exclusion criteria that are listed in Table 2 to be enrolled in the study.

**Table 2.**
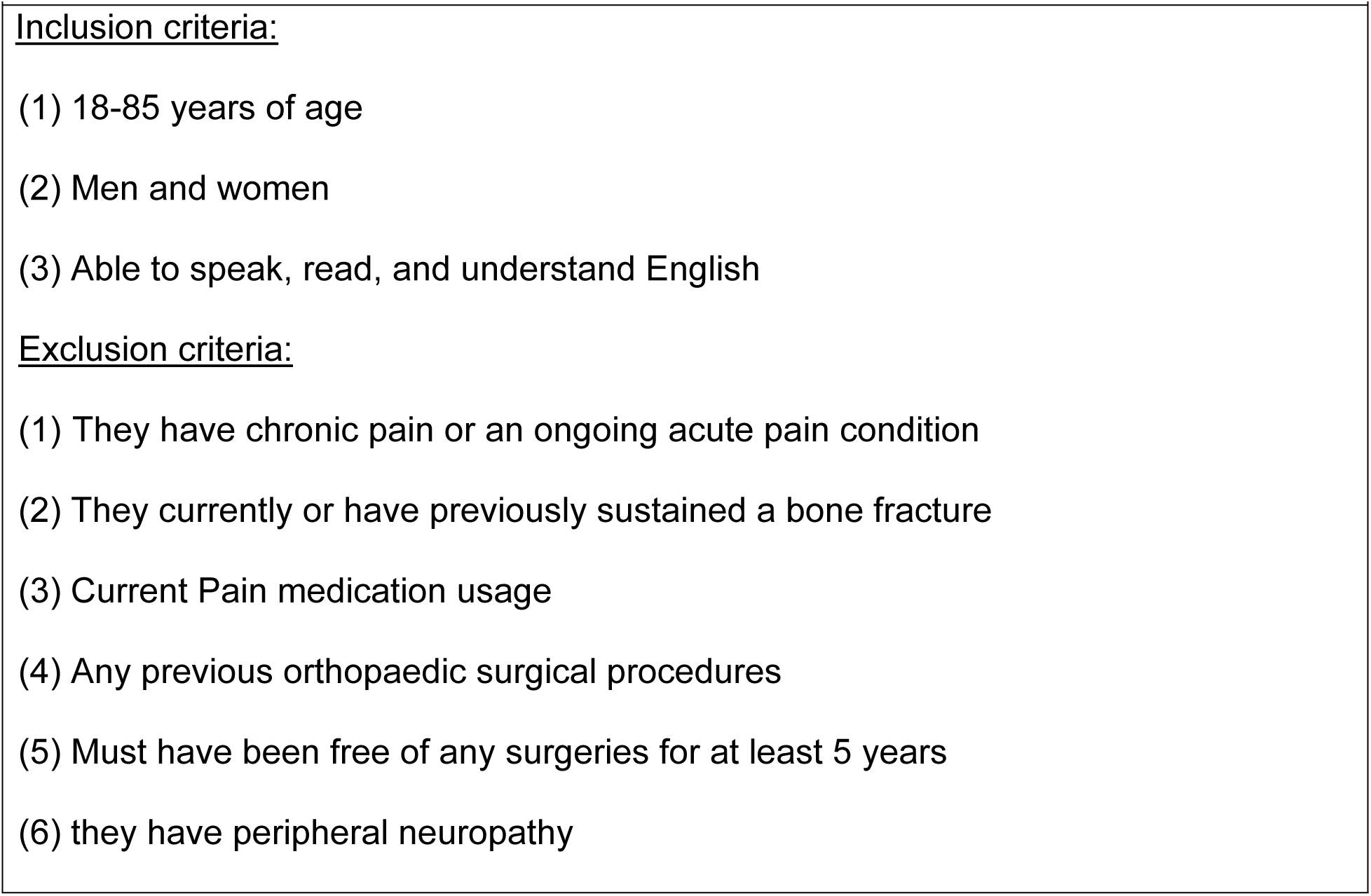
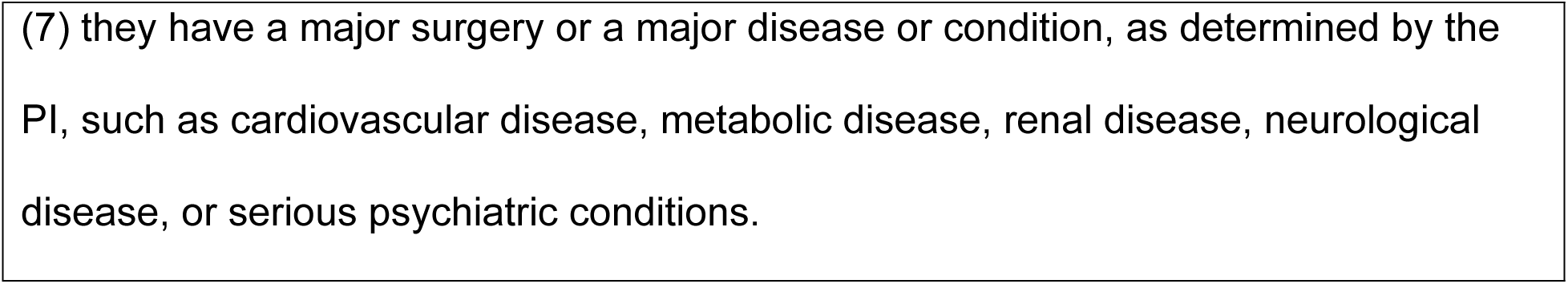
Inclusion and exclusion criteria for controls.

### Recruitment

#### Fracture patients

We will draw our case sample from patients undergoing surgery at IU Methodist Hospital with fractures of the ankle. We have a HIPAA waiver of consent for screening. IU Methodist Hospital Orthopedic research staff will review daily inpatient and outpatient OR schedules to screen for potentially eligible participants. We will be using the hospitals’ electronic medical records to obtain this information. Potentially eligible patients will be called on the phone by a member of the study team after their surgery but before their first post-operative appointment at the outpatient orthopaedic clinic. For those that express initial interest on the phone, study procedures, purpose, and inclusion/exclusion criteria will be reviewed. For those still interested, the patient will be given the following options:

1. provide verbal consent over the phone and then complete several patient- reported outcomes over the phone or online prior to the first in-person visit. A verbal script will be used when consenting subjects over the phone. Also, subjects who consent verbally will be sent the consent form via email to have during the phone call. Written consent is obtained during the first in-person visit.
2. wait to provide any type of consent until the first in-person visit. In this case, all patient-reported outcomes are not completed until the in-person study visit after the written informed consent is obtained.

#### Controls

Our study will recruit control subjects by being listed on the Indiana Clinical and Translational Sciences Institute (CTSI) study listing called the All IN for Health TrialX iConnect. iConnect is a HIPAA-compliant and secure public facing research recruitment platform provided by the Indiana CTSI consisting of a study trial listing and volunteer registry. This system is licensed by Indiana University from TrialX and is monitored by the Indiana University Information Technology Services (UITS) and University Information Security Office (ISO). Study Teams have access to their study listings and participant referrals based on a secure login. This online platform provides researchers with the ability to create a public facing webpage that allows the public to refer themselves to the trial: (https://research.indianactsi.org). The study listings are open to the public and any persons who visit our study page may submit their contact information to the study team through the listing. We will also recruit participants with IRB-approved flyers. Individuals interested in being a control subject will email or call the research staff. For those that express initial interest on the phone/email, study procedures, purpose, and inclusion/exclusion criteria will be reviewed. For those still interested, the individuals will be given the following options:

1. provide verbal consent over the phone and then complete several study related questionnaires over the phone or online prior to their one time in-person visit. A verbal script will be used when consenting subjects over the phone. Also, subjects who consent verbally will be sent the consent form via email to have during the phone call. Written consent is obtained during the in-person visit.
2. wait to provide any type of consent until the in-person visit. In this case, all patient-reported outcomes are not completed until the in-person study visit after the written informed consent is obtained.

### Procedures

See Table 3 for a list and timeline of study procedures for fracture patients. Our patient reported outcomes were chosen to align with the NIH HEAL Initiative (The Helping to End Addiction Long-term) common data elements [21].

**Table 3.** Calendar of Study Procedures for Fracture patients.

| **Table 3. Calendar of Study Procedures for Fracture patients** | **Daily** | **Standard of Care Follow-up** |  |  |  |
| --- | --- | --- | --- | --- | --- |
|  |  | **2-4\*\* weeks** | **5-9\*\* weeks** | **3-5\*\* months** | **6-9\*\* months** |
| Screening | X |  |  |  |  |
| Consent |  | X |  |  |  |
| Inclusion/Exclusion criteria review^ |  | X | X | X | X |
| Patient Characteristics form*** |  | X |  |  |  |
| Medical History form |  | X |  |  |  |
| AIS form |  | X |  |  |  |
| Injury Characteristics form**** |  | X |  |  |  |
| Clinical Follow-up form (fracture complications and healing) ***** |  |  | X | X | X |
| Participant Follow-up form |  |  | X | X | X |
| Thermography |  | X | X | X | X * |
| Blood draw |  | X | X | X | X * |
| Quantitative sensory testing |  |  | X | X | X * |
| Immune cell analysis |  | X | X | X | X * |
| Chronic Pain Grading Scale |  |  |  | X | X |
| DNF – Interview |  |  |  | X | X |
| Ankle OA scale |  |  |  | X | X |
| Pain Map |  | X | X | X | X |
| Foot and Ankle Disability Index |  | X | X | X | X |
| Pain Catastrophizing Scale |  | X | X | X | X |
| PROMIS – Pain intensity |  | X | X | X | X |
| PROMIS – Pain Interference |  | X | X | X | X |
| PROMIS – Pain Behavior |  | X | X | X | X |
| PROMIS – Anxiety |  | X | X | X | X |
| PROMIS – Depression |  | X | X | X | X |
| PROMIS – Physical Function |  | X | X | X | X |
| PROMIS – Neuropathic Pain Quality |  | X | X | X | X |
| PROMIS – Sleep Disturbance |  | X | X | X | X |
| ^Inclusion/Exclusion criteria review will occur prior to patient enrollment via a) medical record review to identify potential fracture participants and b) possibly over the phone. **All patient reported outcomes (PROs) may also be completed over the phone or online at each timepoint, if the patient prefers. *Only performed if session is in person vs. over phone. ***Age, gender, race, education level, socioeconomic status, workman's compensation status. ****Open/Closed fracture classification, fracture description (both classic and AO/OTA classifications). *****Infection, nonunion, post-traumatic arthritis, re-operation. At 1 to 1.5 years, we will review the patient's medical record to assess for complications and we will contact by phone those determined to have BFNP to assess other PROs to see if their clinical pain state has resolved. |  |  |  |  |  |

#### Screening and Enrollment Visit for Fracture patients (1^st^ visit, 2-3 weeks post-surgery)

The patient will complete the written informed consent document for the study at their 1^st^ post-operative standard of care visit with the first study visit completed in conjunction/right after the 1^st^ post-operative standard of care visit. At the first post- operative visit and over the phone (if applicable), study staff will review the informed consent and provide a clear explanation about what the study involves and its risks and benefits. Participants will be encouraged to take sufficient time to ask questions and to consider whether to volunteer to participate. We will also inform subjects about HIPAA regulations. Subjects will be asked to sign an Informed Consent Form to grant authorization for collection of health data. Those individuals who sign the forms will be enrolled in the study. Participant will be provided a written copy of the consent form. The remaining procedures of this visit are listed in Table 1 and further described below.

#### Follow-up visit(s) – Fracture patients only

The first three follow-up visits will be in-person visits and will be completed within the time frame of 5-9 weeks, 3-5 months, and 6-9 months post-ankle fracture surgery.

Lastly, the fourth follow up will be a phone call follow-up at 1-1.5 years post-ankle fracture surgery. The procedures of each visit are listed in Table 1 and further described below.

#### Control subjects study visit

At the in-person study session, study staff will first review the informed consent and provide a clear explanation about what the study involves and its risks and benefits. Participants will be encouraged to take sufficient time to ask questions and to consider whether to volunteer to participate. Subjects will be asked to sign an Informed Consent Form. Those individuals who sign the form will be enrolled in the study. Participants will be provided a written copy of the consent form. All control subjects will only participate in one single study visit. During the visit, controls will complete the following procedures: inclusion/exclusion criteria review, Patient Characteristics form, Medical History form, thermography, blood draw, quantitative sensory testing, pain map, Pain Catastrophizing Scale, and all PROMIS questionnaires except for the Neuropathic Pain Quality questionnaire.

### Outcome measurements

#### Measurement of BFNP in Fracture Patients

The Chronic Pain Grading Scale (CPGS) will be used to determine whether fracture patients have chronic pain in the chronic phase of injury (3-5 month visit and 6-9 month visit). Fracture patients who report persistent pain (non-zero score on the CPGS) at 3-5 months and 6-9 months will also complete the Douleur Neuropathique 4 (DN4) questionnaire at the 3-5 months and 6-9 month visits.

#### Chronic Pain Grading Scale (CPGS)

The CPGS grades chronic pain as a function of pain intensity and pain-related disability reported by the participant for the last three months [22]. The subscale score characteristic pain intensity will be used to categorize participants into those with and without chronic pain [23]. This item is the sum of three questions (current pain, worst pain, average pain over last month), each scored on an 11-point Likert scale with responses ranging from 0 -10 for a maximum of 30 points. The participant will be asked to report only on pain related to the site of fracture. Participants scoring 1 - 30 on the characteristic pain intensity score in the last month at the site of fracture will be classified as having chronic pain. Patients reporting no pain (0) at the site of fracture in the last month will be classified as having no chronic pain. This tool has been used to assess chronic pain following trauma [23–25]. Internal consistency is acceptable with Cronbach’s alpha ranging from 0.67 to 0.74 depending on chronic pain condition and Kappa coefficient from 0.79 to 0.86. The CPGS has also shown good concurrent and prognostic validity [22, 26].

#### Douleur Neuropathique 4 (DN4)

The DN4 is a validated and reliable screening tool for neuropathic pain consisting of 7 items [27, 28]. The items relate to pain quality (i.e., sensory and pain descriptors) and are based on interview with the patient. The items of the DN4 are scored based on a yes (1 point)/no (0 points) answer. This leads to a score range of 0–7. The cut-off value for the classification of neuropathic pain is a total score of >3 [28, 29]. The DN4 has been used to identify neuropathic pain in ankle and wrist facture patients [7, 19].

#### Blood collection for pro-inflammatory mediators

For visits 1-4, a site research nurse will collect up to 450 mL of blood using standard protocols. Blood specimens are considered routine safety laboratories and will include EDTA anticoagulant tubes for plasma analysis and PAXgene for RNA sequencing. This blood will be assayed for the presence of pro-inflammatory mediators.

#### Thermography assessment

For visits 1-4, differences between the two ankles will be measured at room temperature (23°C), with subjects kept in an upright position for 15 min to obtain sympathetic equilibrium. A thermographic image of the ankles will be taken using an infrared camera from a distance of 70 cm. Thermography is a noninvasive tool that uses an infrared camera to produce images that show patterns of heat and blood flow on or near the surface of the body.

#### Quantitative Sensory Tests (QST)

Subjects will be made familiar with each sensory test to be performed and will be taught the 0-100 pain rating system. Subjects are not expected to experience severe pain during the QST procedures as the temperatures and pressures to be used are below the pain tolerance levels observed in human studies. The sensory detection and pain threshold test will be performed first followed by the temporal summation test and then the conditioned pain modulation (CPM) test. Sensory and pain detection thresholds will be measured on the skin proximal to the level of ankle injury to assess neuropathy severity, as well as the contralateral uninjured side to serve as a within subject control. A trained research coordinator will perform the testing. Specifically, the following measures will be administered based on the guidelines provided by the German Research Network in Neuropathic Pain [30]: cold and warm detection and pain thresholds and mechanical detection thresholds. These measures are effective in characterizing small (Aδ, C) and large fiber (Aβ) sensory deficits. We will also administer two of the most extensively studied paradigms to measure endogenous pain modulation in humans: temporal summation (TS) and conditioned pain modulation (CPM), which represent ascending facilitatory and descending inhibitory aspects of central pain processing, respectively [31].

##### Mechanical Detection Thresholds

Mechanical detection thresholds will be examined using von Frey monofilaments, with each filament applied three times in ascending sequence until the threshold is detected in at least two of the three trials. Then, the next lower von Frey filament will be applied, and the lowest filament to be detected at least twice will be determined as the mechanical detection threshold. The subject will close his/her eyes during the test.

##### Thermal detection thresholds and Pain Thresholds

Warm/Cold detection thresholds and heat/cold pain thresholds will be detected using the Q-Sense/Small Fiber Test (Medoc Advanced Medical Systems, Medoc Ltd, Israel). The thermode temperature will increase (heat) or decrease (cold) from a baseline temperature of 32°C with a rise/fall rate of 0.5 °C/s. Participants will be instructed to signal when the stimulus is first perceived as warm/cold. Participants will also be instructed to signal when they first feel the transition from heat to the sensation of heat pain (heat pain threshold) or the transition from cold to the sensation of cold pain (cold pain threshold). Two trials of each will be performed at each ankle.

##### Pressure pain thresholds

A digital, handheld, clinical grade pressure algometer will be used for the mechanical procedures (AlgoMed, Medoc Ltd, Israel). The experimenter will apply a slow constant rate of pressure and record the pressure in kilopascals when the subject responds. Pressure will be applied using a .5 cm^2^ probe. Subjects will be instructed to respond when they first feel pain and that pain threshold will be recorded. Two consecutive measurements at each point with intervals of 20 s will be obtained.

##### Mechanical Temporal Summation of Pain (TS)

TS will be administered on the skin proximal to the level of injury and the contralateral uninjured side to serve as a within subject control. TS can be assessed by administering short-duration repeated noxious stimuli of a constant intensity and measuring the consequent increase in pain as an indirect method of evaluating hyperexcitability of the central nervous system [32]. First, a single pinprick stimulus using a von Frey filament of 6.65 Mn or 300 g will be applied to the body site. Participants will rate the perceived pain intensity using a numeric rating scale of 0 (no pain at all) to 100 (worst pain imaginable). Then, a series of 10 pinprick stimuli (6.65 Mn) will be administered at a rate of 1 Hz, applied to the body site within an area of 1 cm^2^. The temporal summation value will be calculated as the difference between the first and last stimuli. Two TS trials at each body site will be performed.

##### Conditioned Pain Modulation

The most frequently used test of endogenous pain inhibition is condition pain modulation (CPM). CPM refers to the reduction of pain produced by a test stimulus by a second noxious conditioning stimulus in a remote body site (i.e., “pain-inhibition-by-pain”) [33]. CPM will be assessed by determining the ability of a cold-water bath (conditioning stimulus) to diminish pressure pain thresholds (test stimulus) applied at a separate body site. After the first trials of pressure pain thresholds on the left forearm, the contralateral hand will be placed into a cold-water bath for 60s. Then, the second trials of pressure pain thresholds will begin on an adjacent site of the left forearm. *Test stimulus*: Two trials of pressure pain thresholds will be administered on the left volar forearm. A digital, handheld, clinical grade pressure algometer with a .5 cm^2^ probe will be used for the mechanical procedures (AlgoMed, Medoc). The experimenter will apply a slow constant rate of pressure and record the pressure in kilopascals when the subject responds. Subjects will be instructed to respond when they first feel pain and pressure at threshold will be recorded. *Conditioning stimulus*: Participants will immerse their right hand up to the wrist in a cold-water bath maintained at 10°C for up to 1 minute or until they report intolerable pain (we do not expect this to happen because of the moderately cold water used). Cold pain will also be assessed every 15-sec. The dependent variable for the CPM test will be the change in pressure pain threshold of the test stimulus following the conditioning stimulus.

#### Patient reported outcomes

The following questionnaires may be administered on paper, over the phone, or by electronic format (REDCap) via email.

##### Pain Catastrophizing Scale (PCS) – Adult version

The PCS assesses negative mental responses to anticipated or actual pain [34]. The PCS has 13 items that are scored on a Likert scale with three subcategories: rumination, magnification, and helplessness.

Higher PCS scores are indicative of higher pain catastrophizing. Scores on the PCS have been associated with clinical and experimental pain measures [35].

##### Body Pain Map

Participants will locate the areas of the body that they are experiencing pain on an anatomical map of the body [36]. Females will complete a female body map, while males will complete a male body map. The maps present two side-by-side posterior and anterior pictures of the body with a line separating the left and right sides.

##### Ankle Osteoarthritis Scale

The Ankle Osteoarthritis Scale is a reliable and valid self- assessment instrument that specifically measures patient symptoms and disabilities related to ankle arthritis [37].

##### Foot and Ankle Disability Index (FADI)

The FADI is designed to assess functional limitations related to foot and ankle conditions [38]. The FADI contains 26-items, in which each item is scored from 0 (unable to do) to 4 (no difficulty at all). The 4 pain items are scored 0 (unbearable) to 4 (none). This index has excellent reliability and good validity.

##### Patient-Reported Outcome Measurement Information System (PROMIS)

PROMIS is a system of item banks developed by NIH to standardize measurement of patient reported outcomes [39–41]. The PROMIS computer adaptive tests (CAT) will be used to measure pain interference, pain behavior, depression, anxiety, and sleep disturbance. The PROMIS CAT system uses Item Response Theory to develop sample-independent scores that are valid across patients with different demographic characteristics. Other PROMIS tools will be used to measure pain intensity and neuropathic pain quality. The following PROMIS tools will be completed at each visit:

**a. Pain intensity** to measure self-reported current, average, and worst pain in the last seven days on a 5-point scale (none, mild, moderate, severe, or very severe) [39–41].
**b. Pain interference** to measure self-reported effects of pain on social, cognitive, emotional, physical, and recreational aspects of a person’s life from a bank of 40 items [39–41]. The participant rates interference from pain in the last seven days on a 5-point scale (not at all, a little bit, somewhat, quite a bit, very much).
**c. Pain behavior** to measure self-reported external manifestations of pain; behaviors that typically indicate to others that an individual is experiencing pain from a 20-item bank [39–41]. The participant rates their pain behavior in the last seven days on a 6-point scale (no pain, never, rarely, sometimes, often, always).
**d. Sleep disturbance** to measure self-reported perceptions of sleep quality, sleep depth, and restoration associated with sleep from a 27-item bank [39–41]. The participant rates sleep disturbance in the last seven days on a 5-point scale (not at all, a little bit, somewhat, quite a bit, very much).
**e. Depression** to measure self-reported negative mood (sadness, guilt), views of self (self-criticism, worthlessness), social cognition (loneliness, interpersonal alienation), and decreased positive affect and engagement (loss of interest, meaning, and purpose) [39–41]. The participant rates depressive symptoms in the last seven days on a 5-point scale (never, rarely, sometimes, often, always).
**f. Anxiety** to measure self-reported fear (fearfulness, panic), anxious misery (worry, dread), hyperarousal (tension, nervousness, restlessness), and somatic symptoms related to arousal (racing heart, dizziness) from a 29-item bank [42, 43]. Participants rate anxiety in the last seven days on a 5-point scale (never, rarely, sometimes, often, always).
**g. Neuropathic pain quality scale** is a short (5 items) and practical measure that can be used to identify patients more likely to have neuropathic pain and to distinguish levels of neuropathic pain [44, 45].
**h. Physical Function** item bank v1.0 includes a total of 124 physical function items across five categories of physical functioning: upper extremity, lower extremity, axial, central, and instrumental activities of daily living [46]. The items can place patients on the continuum of function from extremely low to very high. Using CAT allows for accuracy similar to delivering all the items, but generally only requires delivery of 4 to 6 items.
**i. Emotional Support** item bank v2.0 measures self-reported perceptions of being emotionally supported by others, including feelings of being loved, cared for, and valued, as well as having someone to talk to during times of need. The participant rates their experiences over the past seven days on a 5-point scale (never, rarely, sometimes, often, always).
**j. Informational Support** item bank v2.0 measures individuals’ perceptions of having access to useful advice, guidance, or information when needed, especially during times of decision-making or problem-solving. Participants rate their experiences on a 5- point scale (never, rarely, sometimes, often, always).
**k. Instrumental Support** item bank v2.0 will be used for self-reported perceptions of receiving practical help from others, such as assistance with chores, errands, or transportation when needed. Participants rate their experiences over the past seven days using a 5-point scale (never, rarely, sometimes, often, always).

#### Data management plan

Primary data will be collected via paper or electronically through REDCap and stored electronically in REDCap and eventually statistical analysis files on a Department Server. The storage location will be backed up automatically every day. Other data sources include blood tubes that will be stored at -80°C. Quality assurance steps will include: 1) built-in range checks and 2) database testing by the study team prior to moving to production mode. The following quality control methods will be used: 1) single entry with random checks of accuracy and 2) extraction and cleaning of data that will be used for analysis every 6 months.

#### Costs and compensation

All eligible fracture participants will receive up to a total of US $300 in gift cards. Specifically, the participants will be paid US $100 Dollars for each completed visit 2-4. Control participants will receive $50 in gift cards. Participants will receive these cards at the end of each testing session. In cases where a participant is unable or wishes not to complete all procedures of the study visits, but are willing to complete partial data collection, such as questionnaires, blood draw, and thermography, they will receive a US $25 gift card for each study visit. In addition, participants who attend in-person visits will also receive a parking validation ticket to cover the cost of parking.

### Statistical analysis

#### Data Analysis

Statistical Analyses/Machine Learning will be conducted under the guidance of our biostatistician. Prior to hypothesis testing, descriptive statistics will be computed for each of the outcome measures, e.g., CPM, TS, and immune biomarkers, by time. The spaghetti plot for each measure over time and distributional characteristics (e.g., normality, outliers, and missingness) will be explored. The proper steps (e.g., transformation) will be taken if needed to ensure the assumptions of the procedure for hypothesis testing are met. To address our aims, we will examine differences in pain modulation (TS, CPM) and immune biomarkers among the fracture participants who develop BFNP and those who do not and healthy controls. We expect fracture patients developing BFNP to exhibit (a) enhanced central sensitization and less efficient endogenous inhibition of pain and (b) alterations in immune cell subsets and cell reactivity post-injury and across time when compared to fracture patients who do not develop chronic pain and matched healthy controls. We will use latent growth mixture models (GMM)[45] to classify the patterns of the trajectory for each measurement by evaluating the latent intercept (baseline means) and latent slope (development) into mutually exclusive classes. GMM will then regress those latent variables to the outcome (BFNP, BF no-chronic pain, and Healthy Control) to test the differences of these factors at 2-3-weeks post injury and across time. Trajectories from the GMM (latent intercept and slopes) will be explained by a nominal latent class variable C. The models will include participant characteristics (e.g., age, race, gender, fracture pattern, etc.). When necessary, a quadratic latent growth variable will be tested to fit the GMM model. The models will be evaluated by the model fit indices such as Akaike information criterion (AIC), Bayesian information criterion (BIC), and likelihood ratio test as well as the corresponding profile plots. GMM is a flexible and more accurate method for measuring change than more traditional approaches such as a univariate repeated measures analyses of variance (ANOVA), a multivariate repeated measures ANOVA, or an analysis of covariance.

#### Missing Data

There will be missing values related to dropout and nonresponse to surveys at different time points. If there is excessive or differential attrition across BFNP, BF no- chronic pain, and Healthy Control groups, we will identify baseline characteristics that differ between participants who did and did not have missing values. We will apply sensitivity analysis, such as inverse probability weighting methods, as needed.[47] If the procedure yields different results for the hypothesis tests, we will report study outcomes both with and without sensitivity analysis. Also, it is possible that participants may be found ineligible after study enrollment. Any data collected from these participants will not be included in the data analysis.

#### Supervised machine learning (ML)

Supervised machine learning predictive models will be implemented using the features such as demographic information, blood biomarkers and QST scores of the study participants. Multiple ML predictive models will be developed and tested to assess their performance and efficacy to better predict which participants will be prone to develop BFNP. The predictive models will be trained/validated and tested using information available 2-3 weeks after surgery and data collected at 6-9 months post-surgery. Our implementation of supervised ML predictive models incorporates these steps: 1) A dataset is gathered. The development dataset needs to be representative of the real-world population and must include reasonably large representations of both BFNP and non-BFNP subjects. 2) Quality controls are applied. All data are subjected to a series of tests to ensure the information is valid and the data do not contain anomalies arising from measurement errors. 3) The features to be used for the prediction models are extracted from the development dataset. The feature extraction will be carried out to transform multiple data types into numerical features usable for ML model training. 4) Feature Selection: Using the domain knowledge and ML based feature selection algorithms, the number of input features will be reduced to decrease computation time and to prevent model overfitting.

A specific ML methodology is selected. We have used a variety of machine learning methodologies including Elastic Net (ENET). ENET is a flexible methodology that, based on how penalty functions are set, can implement various specific methodologies such as Least Absolute Shrinkage, Selection Operator (LASSO) and ridge regression. The algorithm performs both feature selection and regularization to enhance the prediction accuracy and interpretability of the statistical model it produces. Support Vector Machines, K-Nearest Neighbors, Random Forest and Gradient Boost are also used in the methodology on the same training/validation/test dataset, to provide comparison points when establishing the performance of the algorithms generated using ENET. Data split is performed. The model is trained and tested using 70/30 split between training (and validation) and testing datasets. The split is stratified so that training and test subsets have the same proportion of BFNP and non-BFNP class labels as that of the input dataset.

Nested Cross-Validation is performed. Cross-validation provides a way to estimate algorithm performance on an unseen (validation) dataset, using the annotated train/validate dataset in hand. To overcome bias in model performance evaluation, we will use nested cross-validation. We intend to use 10-fold cross-validation (outer-loop) to evaluate model performance and 5-fold cross-validation (inner loop) for model and hyperparameter optimization. In the outer loop 10-fold cross-validation, the original train dataset is randomly portioned into 10 equal-size subsamples. Within the 10 subsamples, a single subsample is considered to be the “validation” group, and the other 9 subsamples (the “train” group) are used for model and hyperparameter optimization. The 10-fold cross-validation procedure involves fitting a model on “train” group and evaluating the fit model on the “validation” group (outer loop). The “train” group is then provided the hyperparameter optimization scheme to find an optimal set of hyperparameters for the model (inner loop). The evaluation of each set of hyperparameters is performed using 5- fold cross-validation that splits up the provided train dataset into 5 folds. In this step (inner loop) we also evaluate settings in terms of their alignment with domain knowledge. The performance of the final model will be assessed on the test data. To estimate variance in model performance and to mitigate selection bias in training/validation/test data, the entire workflow will be repeated 100 times and, during each iteration, the model will be trained with a unique random number of seed. The average of 100 models will be used to estimate model confidence and to extract feature importance. When ML analysis of in-house data reveals patterns in alignment with domain knowledge reported in literature, this indicates the ML-trained algorithm is reflective of underlying phenomena and not overtrained on our dataset.

### Ethical considerations

This study protocol was approved by the IU Institutional Review Board (**IRB 19914**). Informed consent will be obtained from all participants or their legal guardians. All methods are performed in accordance with relevant guidelines and regulations.

Participation in this study is voluntary. Before choosing to participate in the study, participants will be able to discuss any concerns they may have with the study team including time commitment, testing procedures, potential expenses, and side effects. The participants may choose to decline to be part of the study, and they do not have to participate or may withdraw from the study at any time. Leaving the study early will not result in any loss of benefits that they already have.

#### Risks and safety

There are risks to participating in any research study. There is a risk of side effects from the procedures performed during this study. Side effects may vary from person to person. Possible risks include:

##### Blood draw

The risks of drawing blood from a vein include discomfort at the site of puncture; possible bruising and swelling around the puncture site; rarely, an infection; and uncommonly faintness from the procedure. Risks can be reduced by using sterile needles, alcohol scrub, applying pressure to arm after we remove the needle, and using a trained technician experienced in collecting research blood draws.

##### Quantitative sensory testing

During the study session, participants may feel pain. This is intended because this is a pain study. However, in any of the pain tests, participants are not expected to experience severe pain as the temperatures and pressures used are below pain tolerance levels observed in human studies. The pain they experience from these pain tests is temporary and generally diminishes immediately after the pain test. They may also stop any pain test at any time with no adverse consequences. Below is a description of the types of pain testing the participants will experience in this study.

**a. Heat pain from the thermode**: Several things may occur in the skin area (1 inch square) after the heated probe makes contact with the skin: (1) It may turn red like a mild sunburn and (2) there may be a slight burning feeling after the heat is removed. For most people this is gone in 1-2 minutes. It may take up to 1-2 hours for all the symptoms to disappear. In addition, during this thermal sensitivity testing, the participant has a button control that can stop the process at any time if the pain becomes intense.
**b. Cold Pain from the thermode**: When the temperature of the small metal probe touching the skin decreases, the coldness that the subject experiences may become painful and unpleasant. The subject’s skin may turn red for a short period of time. However, the risk associated with this procedure is low because the temperature of the probe will not reach temperatures that will cause blisters or skin freezing.
**c. Pressure/mechanical pain**: The handheld probe that has a small nylon tip is applied just to the point of making a slight indentation without puncturing the skin. When the probe is used to tap the skin ten times in a row, participants may stop this test at any time if it becomes too painful. The pain is temporary, and this procedure will not damage skin. The pressure tool has a flat, rubber tip and will not reach a pressure level to damage tissue. There is a slight chance that a small bruise may form as a result of the pressure pain. If a bruise should appear it is usually short lasting and not painful.
**d. Cold water pain**: While the cold water is perceived as unpleasant, the temperature will be monitored to ensure it remains at appropriate temperatures used successfully in other studies. In addition, the participants can remove their hand(s) from the cold water at any time if the pain becomes too intense.

##### Images of ankle taken with thermal camera

The camera used to take thermal pictures of the participant’s ankle is a non-invasive tool. There is no radiation exposure, no compression of ankles, and no physical risks associated with this procedure.

##### Psychological discomfort

The prospect of being subjected to painful stimulation in an unfamiliar laboratory, surrounded by unfamiliar investigators and equipment that may look intimidating may lead to anxiety. The researchers try to minimize this anxiety by thoroughly explaining all procedures to the participants and taking the time to answer all their questions. In addition, the pain tests always start with a few non-painful stimuli before the temperature or pressure rises gradually. Despite these measures, it is not possible to completely rule out that participants may feel some anxiety during the first session. However, this anxiety should lessen during the second and subsequent sessions, once a participant has become familiar with the experimental procedures.

##### Risk of possible loss of confidentiality

The study team will collect information about the participants from their medical records. This information, some of which may identify the participants, may be used for research-related purposes. This may include making sure the participants meet the criteria to be in this study, gathering information about their medical history to include in the research data, reviewing results of their medical tests for safety purposes, checking on their health in the future to help answer our research question, or to inspect and/or copy their research records for quality assurance and data analysis. There is a risk of possible loss of confidentiality, which means there is a risk someone outside the study team could get access to patient research information from this study. Individuals and organizations including the researchers and research staff on this study, the IU Institutional Review Boards or its designees, the IU Clinical Research Center (ICRC), the US governments or agencies as required by law, the data safety monitoring boards, State or Federal Agencies with research oversight responsibility including but not limited to Office for Human Research Protection (OHRP) and Department of Defense (DOD; Office of Human Research Oversight [OHRO]).

*Biological specimen*, e.g. processed blood sample, will be stored and labeled only with a number identifier. No subject name or personal information will be on this label.

Information and specimens collected for this study may be used for other research studies or shared with other researchers for future research. If this happens, information which could identify the participants, such as names and other identifiers, will be removed before any information or specimens are shared. Since identifying information will be removed, we will not ask for additional consent.

### Status and timeline of the study

The study opened to accrual on January 1^st^, 2024. As of May 31^st^ 2025, we have enrolled 64 fractured participants, 25 of which have completed their 6-9 month study visits, and 6 control participants.

## Discussion

Overall, findings from this longitudinal study will provide a means to identify patients who may be at risk of developing persistent BFNP. Secondly, the profile signatures identified by the multipronged approach in the proposed study will provide important mechanistic insights into understanding factors contributing to the development of BFNP that may help (1) design appropriate treatment paradigms for patients (with currently available pharmacological and non-pharmacological means) and (2) identify appropriate treatment targets (e.g., increasing inhibitory tone and/or decreasing excitability, suppressing immune/inflammatory triggers) for prevention of BFNP after ankle fracture in military and civilian populations.

## Author contributions

Conceptualization: RMN, KMN, FAW

Funding acquisition: RMN, FAW

Methodology: RMN, KMN, TN, FAW

Formal analysis: HAD, MF, RMN, KMN, TN, FAW

Project administration: RMN, FAW

Supervision: RMN, FAW

Writing – original draft: KMN, TN, RMN, FAW

Writing – review and editing: KMN, TN, RMN, FAW

## Data Availability

no data

